# Eccentric Vessel Wall Enhancement and hs-CRP as Prognostic Markers in Acute Ischemic Stroke: A Prospective Cohort Study

**DOI:** 10.1101/2024.06.17.24309035

**Authors:** Seunghee Na, Taewon Kim, Jung-hwi Lee, Jaseong Koo, Yun Jeong Hong, Seong-Hoon Kim

## Abstract

**Background and Purpose:** Eccentric vessel wall enhancement (EVWE) and high-sensitivity C-reactive protein (hs-CRP) are inflammatory biomarkers associated with atherosclerotic disease. We investigated their prognostic value in patients with acute ischemic stroke receiving guideline-adherent medical treatment.

**Methods:** In this prospective observational cohort study, patients with acute non-cardioembolic ischemic stroke underwent vessel wall MRI and hs-CRP testing. The primary outcome was subsequent ischemic stroke during the follow-up period. The median follow-up duration was 21 months. Kaplan-Meier survival and Cox regression analysis was used to determine the associations between EVWE, hs-CRP levels, and subsequent ischemic stroke.

**Results:** Among 191 patients, 81 (42.4%) had EVWE. EVWE positivity showed a trend towards a lower risk of subsequent ischemic stroke compared to EVWE negativity (HR 0.32, 95% CI 0.12-0.87; P=0.061). Hs-CRP levels were not significantly associated with recurrent stroke risk. The combination of EVWE positivity and low hs-CRP levels (<1.25 mg/L) was associated with the most favorable outcome, while EVWE negativity and high hs-CRP levels (≥1.25 mg/L) were associated with the worst outcome (HR 0.143, 95% CI 0.04-0.50; P=0.031).

**Conclusions:** In patients with acute ischemic stroke receiving optimal medical therapy, EVWE positivity may paradoxically indicate a lower risk of recurrent stroke. The combination of EVWE and hs-CRP status provides prognostic information, with EVWE positivity and low hs-CRP levels associated with the most favorable outcome. These findings highlight the potential role of integrating imaging and serum inflammatory biomarkers in risk stratification and management of acute ischemic stroke patients.

## Introduction

Inflammation plays a crucial role in the development of atherosclerosis, a major risk factor for acute ischemic stroke. Recent clinical trials, such as the COLCOT trial and LoDoCo2 trial, have demonstrated the significance of targeting inflammation in reducing cardiovascular events in patients with coronary artery disease.^1,2^ These studies highlight the potential benefits of anti-inflammatory therapies in the prevention and management of atherosclerotic diseases, including ischemic stroke.^1,2^

Intracranial black blood vessel wall magnetic resonance imaging (VW-MRI) has emerged as a valuable tool in detecting inflammatory changes of the intracranial arteries.^3,4^ This imaging technique provides insights into the pathophysiology of acute ischemic stroke and may aid in identifying patients at higher risk for recurrent events. Typical VW-MRI findings of intracranial atherosclerotic disease (ICAD) plaques include focal and eccentric enhancement. ^5–7^ In patients with recent stroke, this enhancement has been reported to be associated with the symptomatic status of the ICAD plaque.^3^ However, the long-term clinical implications of ICAD enhancement remain largely unknown, particularly in the era of advanced medical management, which includes high-potency dual antiplatelet therapy and rigorous management of hypertension, diabetes mellitus, dyslipidemia and cigarette smoking.

High-sensitivity C-reactive protein (hsCRP), an another biomarker marker of inflammation, has been robustly associated with cardiovascular outcomes. A recent collaborative analysis of three randomized trials demonstrated that elevated hsCRP levels were predictive of cardiovascular events, even among patients receiving statin therapy.^8^ This finding underscores the importance of considering inflammatory biomarkers in the risk stratification and management of patients with acute ischemic stroke.

Given the growing evidence supporting the role of inflammation in the pathogenesis and prognosis of cardiovascular events and acute ischemic stroke, the aim of this study is to evaluate the association between radiological (vessel wall enhancement) and serological (hs-CRP) biomarkers of inflammation and long-term cerebrovascular outcomes in patients with acute ischemic stroke. In contrast to most previous cross-sectional studies on vessel wall enhancement, the current prospective study evaluated inflammatory biomarkers and followed up with acute ischemic stroke patients to assess their future cerebrovascular outcomes. By investigating the relationship between these biomarkers and clinical outcomes, we aim to provide insights into the prognostic value of inflammation in the context of acute ischemic stroke and inform future strategies for risk stratification and targeted interventions.

## Methods

### A. Study population and design

This prospective observational cohort study was approved by the Institutional Review Board of The Catholic University of Korea (OC23RISI0149). Upon reasonable request to the corresponding author, aggregate data can be shared. The study adheres to the reporting guideline of the Strengthening the Reporting of Observational Studies in Epidemiology (STROBE) statement. ^9^

We enrolled consecutive patients who admitted to our hospital’s emergency department from November 2021 to October 2022 with acute ischemic stroke. Patients were classified according to the Trial of ORG 10172 in Acute Stroke Treatment (TOAST) stroke classification criteria.^10^ Only patients with large artery atherosclerosis (LAA) or small vessel occlusion (SVO) were included in the study. Patients with other etiologies, such as cardioembolic stroke, and those with other determined or undetermined etiologies (two or more, negative, or incomplete) were excluded from the study. Additionally, patients who received intraarterial endovascular treatment were also excluded to eliminate the potential effect of post-procedural contrast enhancement on VW-MRI results. All participants underwent both vessel wall MRI and high-sensitivity C-reactive protein (hs-CRP) testing. They were then classified into subgroups based on the presence or absence of eccentric vessel wall enhancement (EVWE) or whether their hs-CRP levels were above or below the median value (1.25 mg/L).

Clinical data regarding age, sex, history of hypertension, diabetes mellitus (DM), dyslipidemia, previous ischemic or hemorrhagic stroke, coronary artery disease (CAD), congestive heart failure (CHF), and current cigarette-smoking status were obtained. All patients underwent detailed clinical evaluations, including neurological examinations, laboratory tests, chest radiography, 12-lead electrocardiography, 24-hour Holter monitoring, echocardiography, MRI, and contrast-enhanced MR angiography or computed tomography (CT) angiography from the aortic arch to the intracranial vessels. Demographic, clinical, and neuroimaging data were collected for each participant at enrollment.

### B. Black blood vessel wall MRI

All subjects underwent MRI scans, which included high-spatial resolution three-dimensional MRI with a black blood effect. The scans were acquired using a Siemens MAGNETOM Skyra 3 Tesla MRI scanner with a T1 SPACE (Sampling Perfection with Application optimized Contrasts using different flip angle Evolution) 3D fast spin-echo sequence. The imaging parameters were as follows: sagittal slice thickness, 0.9 mm; repetition time, 500 ms; echo time, 25 ms; flip angle, 120°; voxel size, 0.56 x 0.625 x 0.9 mm.

A neuroradiologist (JH Lee) reviewed the patients’ vascular neuroimaging studies and confirmed the presence of eccentric vessel wall enhancement on VW-MRI in the vessel relevant to the index stroke (Figure 1).

**Figure 1.**
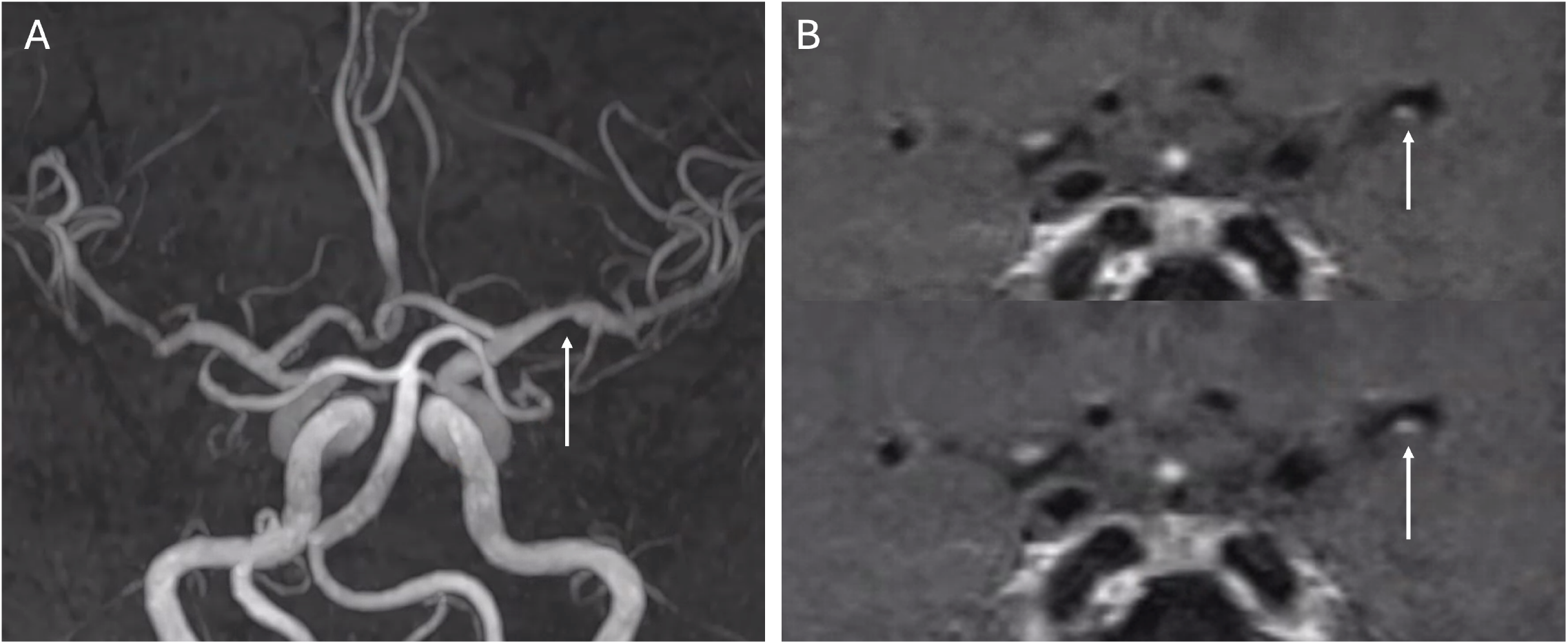
Representative case of eccentric vessel wall enhancement (EVWE) in the left middle cerebral artery of a patient with acute ischemic stroke.

### B. Primary Exposure

In accordance with current stroke guidelines, all patients received aspirin and clopidogrel for at least 21 days, unless antithrombotic therapy was contraindicated due to complications such as active bleeding.^11^ After this period, the decision to maintain monoantiplatelet or dual-antiplatelet therapy was at the discretion of the treating physician. All patients were treated with high-dose statin therapy,^12^ and other vascular risk factors, such as hypertension and diabetes mellitus, were managed according to current guidelines. ^12^

### C. Clinical Outcomes

The primary outcome was a subsequent ischemic stroke during the follow-up period, defined as new or worsening neurological symptoms lasting for at least 24 hours, or lasting less than 24 hours but accompanied by imaging evidence of new or enlarging acute infarction in the territory of the affected artery.

### D. Statistical Analysis

Statistical analyses were performed using SPSS for Windows version 28.0 (IBM Corporation, Armonk, NY, USA). Values are expressed as mean ± standard deviation, median with interquartile range, or percentage.

Baseline characteristics were compared between patients with versus without EVWE using the chi-square test (or Fisher’s exact test) for categorical variables and the independent-samples t-test for continuous variables.

Kaplan-Meier survival analysis was used to assess the subsequent occurrence of ischemic stroke. Unadjusted and adjusted Cox proportional hazards regression models were used to determine the associations between EVWE, hs-CRP levels, and subsequent ischemic stroke. Models were adjusted for prespecified variables known or thought to potentially alter stroke risk and treatment choice, including age, diabetes, hypertension, dyslipidemia, prior stroke, coronary artery disease, active smoking, and chronic kidney disease.

Exploratory subgroup analyses compared ischemic stroke risk between patients with EVWE+ and hs-CRP<1.25 mg/L versus those with EVWE- and hs-CRP≥1.25 mg/L. Baseline characteristics were compared between these subgroups using chi-square tests (or Fisher’s exact test) for categorical variables and t-tests for continuous variables.

Two-sided p-values <0.05 were considered statistically significant.

## Results

### Baseline Characteristics

A total of 191 patients with acute non-cardioembolic ischemic stroke who underwent black blood vessel wall MRI were included in the analysis. Of these, 81 (42.4%) had EVWE+ on MRI, while 110 (57.6%) did not (EVWE-). The baseline demographic and clinical characteristics of the two groups were generally similar (Table 1). No significant differences were observed in age, sex, hypertension, diabetes, dyslipidemia, history of stroke, coronary artery disease, chronic kidney disease, active cigarette smoking, or atrial fibrillation between the EVWE+ and EVWE-groups. The mean and median hs-CRP levels were also comparable between the two groups (P=0.945 and P=0.649, respectively). However, the TOAST classification differed significantly, with a higher proportion of large artery atherosclerosis (LAA) in the EVWE+ group and a higher proportion of small vessel occlusion (SVO) in the EVWE-group (P=0.013).

**Table 1.** Baseline demographic and clinical characteristics of patients with acute ischemic stroke, comparing those with and without eccentric vessel wall enhancement.

|  | Eccentric vessel wall<br>enhancement+<br>(n = 81) | Eccentric vessel wall<br>enhancement -<br>(n = 110) | P-value |
| --- | --- | --- | --- |
| Age, years (±SD) | 69.0 ± 12.7 | 67.9 ± 11.5 | 0.521 |
| Male sex, n (%) | 51 (63.0%) | 68 (61.8%) | 0.881 |
| Hypertension, n (%) | 60 (74.1%) | 80 (72.7%) | 0.870 |
| Diabetes, n (%) | 40 (49.4%) | 45 (40.9%) | 0.244 |
| Dyslipidemia, n (%) | 50 (61.7%) | 53 (48.2%) | 0.078 |
| Stroke, n (%) | 15 (18.5%) | 24 (21.8%) | 0.717 |
| Coronary artery disease, n (%) | 5 (6.2%) | 8 (7.3%) | 1.000 |
| Chronic kidney disease, n (%) | 3 (3.7%) | 4 (3.6%) | 1.000 |
| Active cigarette smoking, n (%) | 26 (32.1%) | 43 (39.1%) | 0.362 |
| *Atrial fibrillation, n (%) | 2 (2.5%) | 1 (0.9%) | 0.575 |
| hs-CRP (mg/L, mean) | 4.4 ± 14.3 | 4.5 ± 21.3 | 0.945 |
| Log<sub>2</sub>hs-CRP | 0.5 ± 1.7 | 0.4 ± 1.8 | 0.649 |
| hs-CRP (mg/L, median, IQR) | 1.25 (0.60-2.68) | 1.23 (0.50-2.55) |  |
| Follow up duration, month (median, IQR) | 20 (3-26) | 22.5 (3-28) |  |
| Treatment of statin | 81 (100%) | 110 (100%) |  |
| Treatment of DAPT | 75 (92.6%) | 105 (95.5%) | 0.532 |
| DAPT duration, days (median, IQR) | 69 (26-140) | 60 (30-128.25) |  |
| DAPT duration, days (mean) | 175.1 ± 280.1 | 119.5 ± 173.2 | 0.117 |
| TOAST classification |  |  |  |
| LAA, n (%) | 50 (61.7%) | 47 (42.7%) | 0.013* |
| SVO, n (%) | 31 (38.3%) | 63 (57.3%) |  |

Abbreviations: hs-CRP, high-sensitivity C-reactive protein; IQR, Interquartile range; DAPT, Dual antiplatelet therapy; TOAST, Trial of ORG 10172 in Acute Stroke Treatment; LAA, large artery atherosclerosis; SVO, small vessel occlusion.
\*Atrial fibrillation: past history or diagnosed during admission.
Values are presented as mean ± standard deviation or number (%). Chi-square test or Fisher's exact test was used for categorical variables, and independent-samples t-test was used for continuous variables.

All patients in both the EVWE+ and EVWE-groups received statin therapy (100% in each group). DAPT treatment was administered to 75 (92.6%) patients in the EVWE+ group and 105 (95.5%) patients in the EVWE-group, with no significant difference between the groups (P=0.532). The median duration of DAPT was 69 days (IQR 26-140) in the EVWE+ group and 60 days (IQR 30-128.25) in the EVWE-group.

A supplementary analysis was conducted to directly compare the baseline characteristics of patients with EVWE+ and hs-CRP<1.25 mg/L (n=40) to those with EVWE- and hs-CRP≥1.25 mg/L (n=55) (Supplementary Table). No significant differences were found in age, sex, hypertension, diabetes, dyslipidemia, history of stroke, coronary artery disease, chronic kidney disease, active cigarette smoking, atrial fibrillation, follow-up duration, DAPT treatment, or DAPT duration between the two groups (all P>0.05). The TOAST classification also did not differ significantly between the groups (P=0.094).

### Association between EVWE, hs-CRP, and Ischemic Stroke

Kaplan-Meier survival analysis showed a trend towards a lower risk of subsequent ischemic stroke in the EVWE+ group compared to the EVWE-group, although this did not reach statistical significance (HR 0.32, 95% CI 0.12-0.87; P=0.061) (Figure 2). When stratified by hs-CRP levels, patients with hs-CRP≥1.25 mg/L had a higher risk of subsequent ischemic stroke compared to those with hs-CRP<1.25 mg/L, but this association was also not statistically significant (HR 2.31, 95% CI 0.87-6.16; P=0.108).

**Figure 2.**
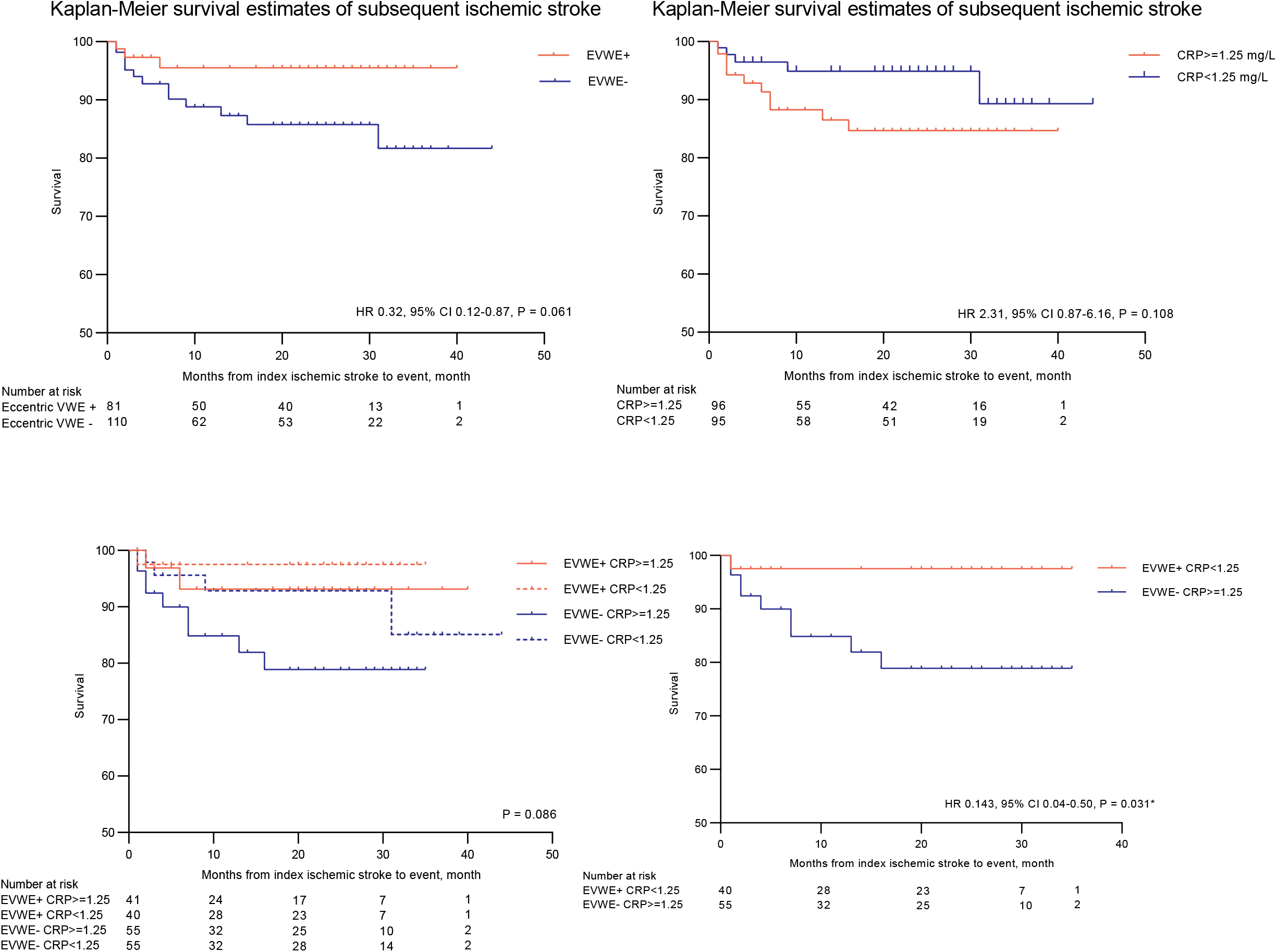
Kaplan-Meier survival estimates of subsequent ischemic stroke based on eccentric vessel wall enhancement (EVWE) status, C-reactive protein (CRP) levels, and their combination. (A) Patients with EVWE+ showed a trend towards a lower risk of subsequent ischemic stroke compared to those with EVWE-, although this did not reach statistical significance (HR 0.32, 95% CI 0.12-0.87; P = 0.061). (B) Patients with CRP levels ≥1.25 mg/L had a higher risk of subsequent ischemic stroke compared to those with CRP levels <1.25 mg/L, but this association was not statistically significant (HR 2.31, 95% CI 0.87-6.16; P = 0.108). (C) Kaplan-Meier survival estimates of subsequent ischemic stroke based on the combination of EVWE status and CRP levels (P = 0.086). (D) Patients with EVWE+ and CRP levels <1.25 mg/L had a significantly lower risk of subsequent ischemic stroke compared to those with EVWE- and CRP levels ≥1.25 mg/L (HR 0.143, 95% CI 0.04-0.50; P = 0.031).

The combination of EVWE and hs-CRP status was further analyzed. Patients with EVWE+ and hs-CRP<1.25 mg/L had a significantly lower risk of subsequent ischemic stroke compared to those with EVWE- and hs-CRP≥1.25 mg/L (HR 0.143, 95% CI 0.04-0.50; P=0.031).

Cox regression analysis was performed to assess the association between EVWE, hs-CRP levels, and ischemic stroke (Table 2). In the unadjusted model, EVWE+ showed a trend towards a lower risk of ischemic stroke compared to EVWE-(HR 0.32, 95% CI 0.09-1.13; P=0.076). Hs-CRP levels ≥1.25 mg/L and ≥2 mg/L were not significantly associated with ischemic stroke risk (P=0.120 and P=0.410, respectively). The combination of EVWE+ and hs-CRP<1.25 mg/L was associated with a lower risk of ischemic stroke compared to EVWE- and hs-CRP≥1.25 mg/L, although this did not reach statistical significance in the unadjusted model (HR 0.14, 95% CI 0.02-1.13; P=0.065).

**Table 2.** Association between eccentric vessel wall enhancement, C-reactive protein levels, and subsequent ischemic stroke: unadjusted and adjusted Cox regression analysis.

|  | Unadjusted<br>HR (95% CI); *P* value | *Model 1<br>HR (95% CI); *P* value | †Model 2<br>HR (95% CI); *P* value |
| --- | --- | --- | --- |
| EVWE+<br>(vs. EVWE- (ref)) | 0.32 (0.09 – 1.13);<br>*P* = 0.076 | 0.32 (0.09 – 1.12);<br>*P* = 0.075 | 0.32 (0.09 – 1.15);<br>*P* = 0.080 |
| hs-CRP ≥ 1.25<br>(vs. <1.25 mg/L (ref)) | 2.31 (0.80 – 6.66);<br>*P* = 0.120 | 2.11 (0.71 – 6.26);<br>*P* = 0.179 | 2.05 (0.68 – 6.21);<br>*P* = 0.206 |
| hs-CRP ≥ 2<br>(vs. <2 mg/L (ref)) | 1.51 (0.56 – 4.07);<br>*P* = 0.410 | 1.40 (0.51 – 3.83);<br>*P* = 0.518 | 1.26 (0.45 – 3.51);<br>*P* = 0.664 |
| EVWE+ & CRP<1.25 mg/L<br>(vs. EVWE- & CRP≥1.25<br>mg/L) | 0.14 (0.02 – 1.13);<br>*P* = 0.065 | 0.14 (0.02 – 1.12);<br>*P* = 0.063 | 0.13 (0.02 – 1.15);<br>*P* = 0.067 |

Abbreviations: EVWE, eccentric vessel wall enhancement; hs-CRP, high-sensitivity C-reactive protein.
\*Model 1: adjusted by age and diabetes mellitus
†Model 2: Model 1 + adjusted by hypertension, dyslipidemia, prior stroke, CAD, active cigarette smoking and CKD.

After adjusting for age and diabetes (Model 1) and further adjusting for hypertension, dyslipidemia, stroke, coronary artery disease, active cigarette smoking, and chronic kidney disease (Model 2), the associations remained similar, with HRs for EVWE+, hs-CRP≥1.25 mg/L, and hs-CRP≥2 mg/L of 0.32 (95% CI 0.09-1.15; P=0.080), 2.05 (95% CI 0.68-6.21; P=0.206), and 1.26 (95% CI 0.45-3.51; P=0.664), respectively. The combination of EVWE+ and hs-CRP<1.25 mg/L remained associated with a lower risk of ischemic stroke compared to EVWE- and hs-CRP≥1.25 mg/L in both adjusted models, although not statistically significant (Model 1: HR 0.14, 95% CI 0.02-1.12; P=0.063; Model 2: HR 0.13, 95% CI 0.02-1.15; P=0.067).

## Discussion

In this study, we investigated the association between two inflammatory biomarkers, EVWE on MRI and serum hs-CRP levels, and the risk of subsequent ischemic stroke in patients with acute non-cardioembolic ischemic stroke who received guideline-adherent medical treatment. Our results suggest that the presence of EVWE, paradoxically, showed a trend towards a lower risk of recurrent ischemic stroke, although this did not reach statistical significance. In our study population, hs-CRP levels were neither associated with EVWE nor with an increased risk of subsequent ischemic stroke. However, the combination of EVWE positivity and low hs-CRP levels was associated with a favorable outcome compared to the combination of EVWE negativity and high hs-CRP levels, a finding that was statistically significant in the Kaplan-Meier survival analysis.

Previous studies have reported that EVWE on high-resolution vessel wall MRI is associated with the symptomatic status of ICAD plaques in patients with recent stroke. ^4,6,7^ The underlying pathophysiologic mechanism of EVWE is thought to involve the accumulation of gadolinium-based contrast agents in the neovascularization and increased endothelial permeability of the atherosclerotic plaque, which are features of plaque vulnerability and inflammation. ^13,14^ These findings suggest that EVWE may be a marker of plaque instability and increased risk of future cerebrovascular events. In addition to its prognostic value, EVWE has been shown to have diagnostic utility in differentiating ICAD from other intracranial vasculopathies, such as moyamoya disease, vasculitis, and reversible cerebral vasoconstriction syndrome. ^6,15^ Thus, the presence of EVWE may guide the selection of patients who could benefit from more aggressive antithrombotic therapy or novel anti-inflammatory treatments. Furthermore, our study found that patients with EVWE had a trend towards a lower risk of subsequent ischemic stroke when treated with intensive medical therapy, including dual antiplatelet therapy, high-dose statins, and optimal management of hypertension and diabetes mellitus. This finding suggests that EVWE may identify patients who are more likely to benefit from aggressive risk factor modification and antiplatelet therapy, potentially leading to a paradoxically favorable outcome. Our results highlight the importance of considering the interaction between imaging biomarkers and treatment strategies when assessing the prognostic value of EVWE in patients with acute ischemic stroke.

Regarding hs-CRP, previous studies have demonstrated that elevated hs-CRP levels are associated with an increased risk of cardiovascular events, even among patients receiving statin therapy. ^8^ Hs-CRP is an acute-phase reactant produced by the liver in response to inflammatory cytokines, such as interleukin-6 and tumor necrosis factor-alpha, which are released in the context of atherosclerosis and other inflammatory conditions. ^16^ Elevated hs-CRP levels reflect the presence of systemic inflammation and have been shown to predict the risk of future cardiovascular events, including ischemic stroke, independently of traditional risk factors. ^17,18^ Moreover, hs-CRP has been implicated in the pathogenesis of atherosclerosis through various mechanisms, such as endothelial dysfunction, monocyte recruitment, and platelet activation. ^19,20^ These findings suggest that hs-CRP is not merely a biomarker of inflammation but may also actively contribute to the development and progression of atherosclerotic disease. However, in our study, hs-CRP levels were not significantly associated with an increased risk of subsequent ischemic stroke. This finding is in contrast with the results of previous studies and may be attributed to several factors. First, our study population consisted of patients who received optimal medical management, including high-dose statin therapy, which has been shown to reduce hs-CRP levels and may have attenuated the association between hs-CRP and recurrent stroke risk. ^21^ Second, the relatively small sample size and low number of recurrent stroke events in our study may have limited our ability to detect a significant association between hs-CRP and clinical outcomes.

Despite the lack of a significant association between hs-CRP and recurrent stroke risk in our study, we found that the combination of EVWE and hs-CRP status provided prognostic information. Patients with EVWE positivity and low hs-CRP levels had the most favorable outcome, while those with EVWE negativity and high hs-CRP levels had the worst outcome. This finding suggests that the integration of imaging and serum biomarkers of inflammation may offer a more comprehensive assessment of risk in patients with acute ischemic stroke.

Interestingly, contrary to our expectation, EVWE was not associated with hs-CRP levels in our study population. This finding suggests that EVWE and hs-CRP may reflect different aspects of the inflammatory process in the pathogenesis of ischemic stroke. One possible explanation for the lack of association between EVWE and hs-CRP is that EVWE may represent a localized inflammatory response within the intracranial vessel wall, while hs-CRP reflects systemic inflammation. ^22^ The presence of EVWE may indicate the vulnerability of an atherosclerotic plaque, which can be influenced by local factors such as shear stress, endothelial dysfunction, and the accumulation of inflammatory cells. ^23^ In contrast, hs-CRP levels are determined by the overall inflammatory burden in the body, which can be affected by various conditions, including obesity, smoking, and chronic infections. ^24^ Therefore, the dissociation between EVWE and hs-CRP suggests that local and systemic inflammation may have distinct roles in the development and progression of ischemic stroke.

The clinical implication of this finding is that the combined assessment of EVWE and hs-CRP may provide a more comprehensive evaluation of the inflammatory status in patients with acute ischemic stroke. The presence of EVWE, even in the absence of elevated hs-CRP, may identify patients with vulnerable intracranial plaques who could benefit from targeted interventions, such as intensive antithrombotic therapy or plaque-stabilizing agents. ^25^ Conversely, elevated hs-CRP levels, regardless of EVWE status, may indicate the need for systemic anti-inflammatory treatments to reduce the risk of recurrent stroke and other cardiovascular events. ^26^ Further research is needed to elucidate the complex interplay between local and systemic inflammation in the pathogenesis of ischemic stroke and to develop personalized treatment strategies based on the individual inflammatory profile.

Our study has several limitations. First, the sample size was relatively small, and the number of recurrent ischemic stroke events was low, which may have limited our ability to detect statistically significant associations between the inflammatory biomarkers and clinical outcomes. Second, we did not assess the dynamic changes in EVWE and hs-CRP levels over time, which may provide additional prognostic information. Third, all participants in our study received the most optimal medical treatment, which may have influenced the observed associations between the inflammatory biomarkers and clinical outcomes. As a result, we could not determine the pure effect of EVWE or hs-CRP on the risk of recurrent ischemic stroke in the absence of medical treatment.

In conclusion, our study suggests that EVWE positivity may identify patients with acute ischemic stroke who are more likely to benefit from intensive medical therapy, leading to a paradoxically favorable outcome. Contrary to our expectation and previous studies, hs-CRP levels were not significantly associated with an increased risk of recurrent ischemic stroke in our population. However, the combination of EVWE and hs-CRP status provided prognostic information, with patients exhibiting EVWE positivity and low hs-CRP levels having the most favorable outcome. These findings underscore the potential value of integrating imaging and serum biomarkers of inflammation in the risk stratification and management of patients with acute ischemic stroke. Further large-scale, prospective studies are warranted to validate these results and explore the role of personalized treatment strategies based on the individual inflammatory profile.

## Data Availability

Upon reasonable request to the corresponding author, aggregate data can be shared

## Acknowledgements

None

## Nonstandard Abbreviations and Acronyms

CAD: coronary artery disease
CHF: congestive heart failure
CT: computed tomography
DM: diabetes mellitus
EVWE: eccentric vessel wall enhancement
Hs-CRP: High-sensitivity C-reactive protein
ICAD: intracranial atherosclerotic disease
LAA: large artery atherosclerosis
MRI: magnetic resonance image
SPACE: Sampling Perfection with Application optimized Contrasts using different flip angle Evolution
SVO: small vessel occlusion
TOAST: the trial of ORG 10172 in acute stroke treatment
VW-MRI: vessel wall magnetic resonance imaging

## References

1. Nidorf SM, Fiolet ATL, Mosterd A, Eikelboom JW, Schut A, Opstal TSJ, The SHK, Xu XF, Ireland MA, Lenderink T, et al. Colchicine in Patients with Chronic Coronary Disease. The New England journal of medicine. 2020;383:1838–1847. doi: 10.1056/NEJMoa2021372

2. Tardif JC, Kouz S, Waters DD, Bertrand OF, Diaz R, Maggioni AP, Pinto FJ, Ibrahim R, Gamra H, Kiwan GS, et al. Efficacy and Safety of Low-Dose Colchicine after Myocardial Infarction. The New England journal of medicine. 2019;381:2497–2505. doi: 10.1056/NEJMoa1912388

3. Edjlali M, Qiao Y, Boulouis G, Menjot N, Saba L, Wasserman BA, Romero JM. Vessel wall MR imaging for the detection of intracranial inflammatory vasculopathies. Cardiovasc Diagn Ther. 2020;10:1108–1119. doi: 10.21037/cdt-20-324

4. Mandell DM, Mossa-Basha M, Qiao Y, Hess CP, Hui F, Matouk C, Johnson MH, Daemen MJ, Vossough A, Edjlali M, et al. Intracranial Vessel Wall MRI: Principles and Expert Consensus Recommendations of the American Society of Neuroradiology. AJNR American journal of neuroradiology. 2017;38:218–229. doi: 10.3174/ajnr.A4893

5. Kwee RM, Qiao Y, Liu L, Zeiler SR, Wasserman BA. Temporal course and implications of intracranial atherosclerotic plaque enhancement on high-resolution vessel wall MRI. Neuroradiology. 2019;61:651–657. doi: 10.1007/s00234-019-02190-4

6. Mossa-Basha M, Shibata DK, Hallam DK, de Havenon A, Hippe DS, Becker KJ, Tirschwell DL, Hatsukami T, Balu N, Yuan C. Added Value of Vessel Wall Magnetic Resonance Imaging for Differentiation of Nonocclusive Intracranial Vasculopathies. Stroke. 2017;48:3026–3033. doi: 10.1161/strokeaha.117.018227

7. Qiao Y, Zeiler SR, Mirbagheri S, Leigh R, Urrutia V, Wityk R, Wasserman BA. Intracranial plaque enhancement in patients with cerebrovascular events on high-spatial-resolution MR images. Radiology. 2014;271:534–542. doi: 10.1148/radiol.13122812

8. Ridker PM, Bhatt DL, Pradhan AD, Glynn RJ, MacFadyen JG, Nissen SE. Inflammation and cholesterol as predictors of cardiovascular events among patients receiving statin therapy: a collaborative analysis of three randomised trials. Lancet. 2023;401:1293–1301. doi: 10.1016/s0140-6736(23)00215-5

9. von Elm E, Altman DG, Egger M, Pocock SJ, Gøtzsche PC, Vandenbroucke JP. Strengthening the Reporting of Observational Studies in Epidemiology (STROBE) statement: guidelines for reporting observational studies. BMJ (Clinical research ed). 2007;335:806–808. doi: 10.1136/bmj.39335.541782.AD

10. Adams HP, Jr., Bendixen BH, Kappelle LJ, Biller J, Love BB, Gordon DL, Marsh EE, 3rd. Classification of subtype of acute ischemic stroke. Definitions for use in a multicenter clinical trial. TOAST. Trial of Org 10172 in Acute Stroke Treatment. Stroke. 1993;24:35–41. doi: 10.1161/01.str.24.1.35

11. Powers WJ, Rabinstein AA, Ackerson T, Adeoye OM, Bambakidis NC, Becker K, Biller J, Brown M, Demaerschalk BM, Hoh B, et al. Guidelines for the Early Management of Patients With Acute Ischemic Stroke: 2019 Update to the 2018 Guidelines for the Early Management of Acute Ischemic Stroke: A Guideline for Healthcare Professionals From the American Heart Association/American Stroke Association. Stroke. 2019;50:e344–e418. doi: 10.1161/str.0000000000000211

12. Kleindorfer DO, Towfighi A, Chaturvedi S, Cockroft KM, Gutierrez J, Lombardi-Hill D, Kamel H, Kernan WN, Kittner SJ, Leira EC, et al. 2021 Guideline for the Prevention of Stroke in Patients With Stroke and Transient Ischemic Attack: A Guideline From the American Heart Association/American Stroke Association. Stroke. 2021;52:e364–e467. doi: 10.1161/str.0000000000000375

13. Choi YJ, Jung SC, Lee DH. Vessel Wall Imaging of the Intracranial and Cervical Carotid Arteries. J Stroke. 2015;17:238–255. doi: 10.5853/jos.2015.17.3.238

14. Portanova A, Hakakian N, Mikulis DJ, Virmani R, Abdalla WM, Wasserman BA. Intracranial vasa vasorum: insights and implications for imaging. Radiology. 2013;267:667–679. doi: 10.1148/radiol.13112310

15. Mossa-Basha M, Alexander M, Gaddikeri S, Yuan C, Gandhi D. Vessel wall imaging for intracranial vascular disease evaluation. J Neurointerv Surg. 2016;8:1154–1159. doi: 10.1136/neurintsurg-2015-012127

16. Pepys MB, Hirschfield GM. C-reactive protein: a critical update. J Clin Invest. 2003;111:1805–1812. doi: 10.1172/jci18921

17. Kaptoge S, Di Angelantonio E, Lowe G, Pepys MB, Thompson SG, Collins R, Danesh J. C-reactive protein concentration and risk of coronary heart disease, stroke, and mortality: an individual participant meta-analysis. Lancet. 2010;375:132–140. doi: 10.1016/s0140-6736(09)61717-7

18. Ridker PM, Rifai N, Rose L, Buring JE, Cook NR. Comparison of C-reactive protein and low-density lipoprotein cholesterol levels in the prediction of first cardiovascular events. The New England journal of medicine. 2002;347:1557–1565. doi: 10.1056/NEJMoa021993

19. Fay WP. Linking inflammation and thrombosis: Role of C-reactive protein. World J Cardiol. 2010;2:365–369. doi: 10.4330/wjc.v2.i11.365

20. Pasceri V, Willerson JT, Yeh ET. Direct proinflammatory effect of C-reactive protein on human endothelial cells. Circulation. 2000;102:2165–2168. doi: 10.1161/01.cir.102.18.2165

21. Ridker PM, Danielson E, Fonseca FA, Genest J, Gotto AM, Jr., Kastelein JJ, Koenig W, Libby P, Lorenzatti AJ, MacFadyen JG, et al. Rosuvastatin to prevent vascular events in men and women with elevated C-reactive protein. The New England journal of medicine. 2008;359:2195–2207. doi: 10.1056/NEJMoa0807646

22. Gupta A, Baradaran H, Al-Dasuqi K, Knight-Greenfield A, Giambrone AE, Delgado D, Wright D, Teng Z, Min JK, Navi BB, et al. Gadolinium Enhancement in Intracranial Atherosclerotic Plaque and Ischemic Stroke: A Systematic Review and Meta-Analysis. J Am Heart Assoc. 2016;5. doi: 10.1161/jaha.116.003816

23. Tuenter A, Selwaness M, Arias Lorza A, Schuurbiers JCH, Speelman L, Cibis M, van der Lugt A, de Bruijne M, van der Steen AFW, Franco OH, et al. High shear stress relates to intraplaque haemorrhage in asymptomatic carotid plaques. Atherosclerosis. 2016;251:348–354. doi: 10.1016/j.atherosclerosis.2016.05.018

24. Hage FG. C-reactive protein and hypertension. J Hum Hypertens. 2014;28:410–415. doi: 10.1038/jhh.2013.111

25. Chung JW, Hwang J, Lee MJ, Cha J, Bang OY. Previous Statin Use and High-Resolution Magnetic Resonance Imaging Characteristics of Intracranial Atherosclerotic Plaque: The Intensive Statin Treatment in Acute Ischemic Stroke Patients With Intracranial Atherosclerosis Study. Stroke. 2016;47:1789–1796. doi: 10.1161/strokeaha.116.013495

26. Ridker PM, Everett BM, Thuren T, MacFadyen JG, Chang WH, Ballantyne C, Fonseca F, Nicolau J, Koenig W, Anker SD, et al. Antiinflammatory Therapy with Canakinumab for Atherosclerotic Disease. The New England journal of medicine. 2017;377:1119–1131. doi: 10.1056/NEJMoa1707914

